# State-level variation of initial COVID-19 dynamics in the United States: The role of local government interventions

**DOI:** 10.1101/2020.04.14.20065318

**Authors:** Easton R. White, Laurent Hébert-Dufresne

## Abstract

During an epidemic, metrics such as *R*_0_, doubling time, and case fatality rates are important in understanding and predicting the course of an epidemic. However, if collected over country or regional scales, these metrics hide important smaller-scale, local dynamics. We examine how commonly used epidemiological metrics differ for each individual state within the United States during the initial COVID-19 outbreak. We found that the case number, and trajectory of cases, differs considerably between states. We show that early non-pharmaceutical, government actions, were the most important determinant of epidemic dynamics. In particular, restricting restaurant operations was correlated with increased doubling times. Although individual states are clearly not independent, they can serve as small, natural experiments in how different demographic patterns and government responses can impact the course of an epidemic.

Daily updates to figures in this manuscript are available at: https://github.com/eastonwhite/COVID19_US_States

## Introduction

The global COVID-19 (caused by the SARS-CoV-2 virus) outbreak began in Wuhan, China in late 2019 (WHO 2020). As of April 12^th^, 1,846,679 cases have been reported across 185 countries and regions. There have been several sets of efforts to track the progression of the outbreak across the world and within countries. For example, John Hopkins University Center for Systems Science and Engineering (CSSE) has compiled data from various sources, including the US Center for Disease Control and the World Health Organization, to present a global picture of COVID-19 cases and deaths (Dong *et al*. 2020). These efforts have allowed for international scientific research and political decision-making. Although data are collected at local scales (e.g. within hospitals), in an emerging pandemic, data are typically reported at the country level. This allows for interesting comparisons between countries (Anderson *et al*. 2020, Jombart *et al*. 2020) and for information from an earlier affected country to be used to slow the outbreak in other places. For instance, South Korea was able to “flatten their outbreak curve” through early and widespread testing as well as strict quarantine policies (Utsunomiya *et al*. 2020). However, country-level analyses still hide more local dynamics that are important to the overall epidemic progression (Chin *et al*. 2020, Lin *et al*. 2020).

Spatial heterogeneity is important for population dynamics generally (Levin 1992, Hanski 2001, Schreiber 2010) and in particular for understanding the progression of infectious disease dynamics (Grenfell *et al*. 1995, Park 2012). Spatial heterogeneity can include differences in local population density, movement patterns, suitability of environmental conditions for transmission, among other factors (Grenfell *et al*. 1995, Park 2012, St-Onge *et al*. 2020). For instance, Keeling *et al*. (2001) showed how spatial distribution and size of farms affected the 2001 UK Foot and Mouth Epidemic.

Here we provide a descriptive analysis of the reported progression of COVID-19 at the state level within the United States. We examine how commonly-used metrics, focusing on doubling time, can vary by state. Clearly, controlled and randomized experiments of COVID-19 spread are not possible (White *et al*. 2019). Therefore, although states are not independent units, we can use state-level data to understand the progression of the outbreak across different replicates within a country (Adolph *et al*. 2020). We show that across measures of demography, education, health, and wealth, only population density was correlated with doubling time. Further, we show that doubling time was more tightly correlated with state-level governmental actions, including restricting businesses.

## Results and Discussion

We used data compiled by John Hopkins University Center for Systems Science and Engineering (Dong *et al*. 2020). The United States has seen exponential growth in the number of cases, especially since February 29th (Fig. S1). Country-level results, however, hide underlying dynamics within each state (Chin *et al*. 2020, Lin *et al*. 2020).

Therefore, we examined how the number of cases changed over time within each state. To properly compare the progression of the epidemic across states, we looked at the log number of cases since the first day a state reported 25 (after which exponential curves were more reliable) cases (Fig. 1). On a log scale, a straight line of the cases over time indicates exponential growth where the slope of the line is the exponential growth parameter.

**Figure 1:**
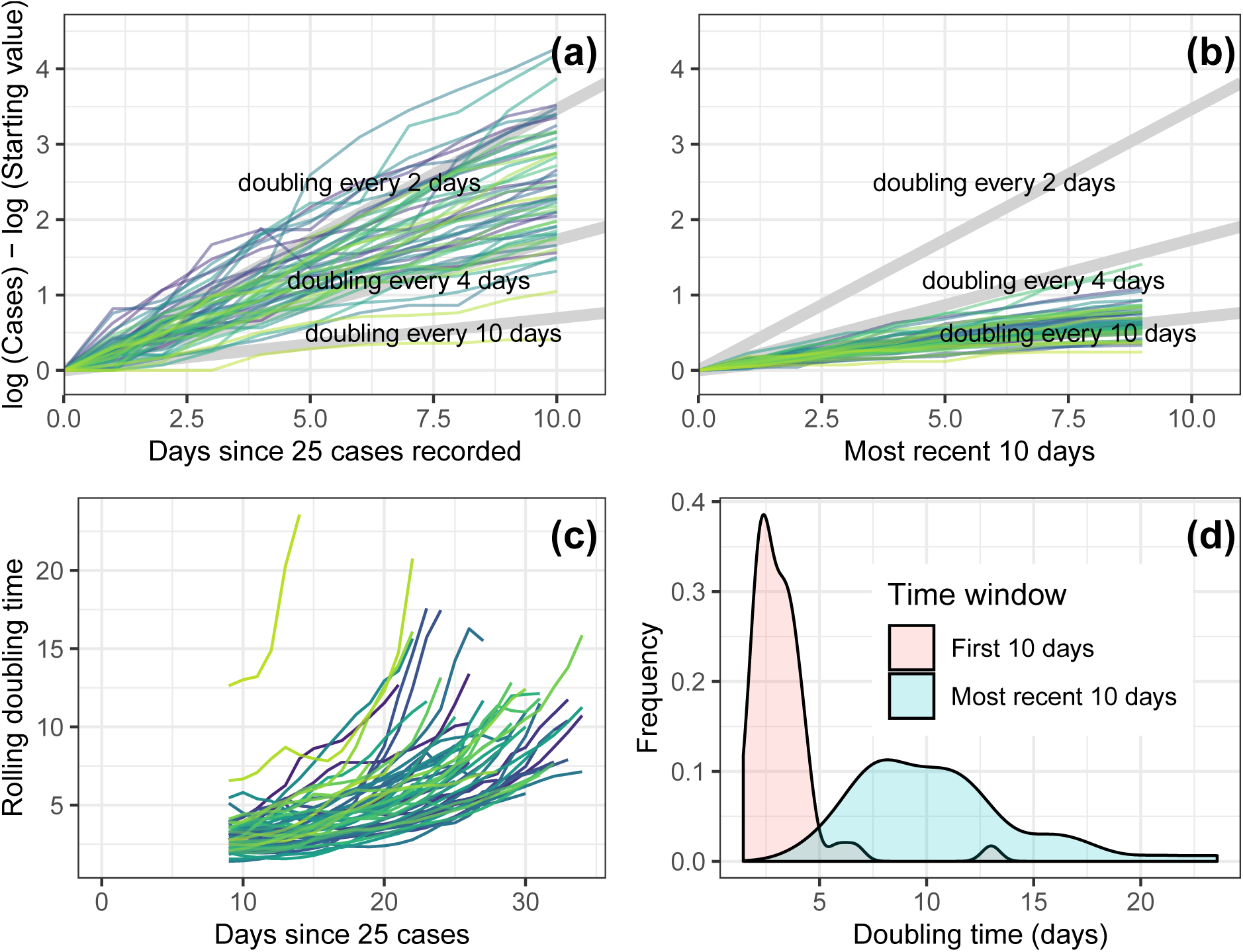
(a) The log number of cases over time for each individual state for the 10 days since their first day of 25 total cases. (b) The log number of cases over time for each individual state for the most recent 10 days. The light grey diagonal lines represent the growth trajectory for doubling times of 2, 4, and 10 days. The log number of the starting value (intial number of cases on first day when at least 25 cases were recorded) had to be subtracted on the y-axis to standarize the graph across states. (c) Rolling doubling times calculated over 10-day windows for each individual states. (d) Distributions of state-level doubling times early and more recent in the course of the outbreak. The figure was produced on 14-Apr-2020.

### State-level variation in COVID-19 trajectories

We found considerable differences between states in how the outbreak has progressed (Fig. 1). These doubling times are, of course, changing over time. We found that doubling times for all states did increase with time and that the heterogeneity between states was reduced (Fig. 1). We mapped doubling time across the US and found regional differences where the West and Northeast have seen large doubling times, i.e. slower outbreak dynamics (Fig. S3).

### Predictors of overall state-level trajectories

Each US state varies considerably across a number of important axes: wealth, access to healthcare, number of international travelers, age distribution, population density, among other factors (Chin *et al*. 2020). In addition, much of the response to COVID-19 has been done at the state, as opposed to federal, government level in the US (Adolph *et al*. 2020, Gostin *et al*. 2020). We examined three hypotheses to explain the state-level variation in COVID-19 trajectories: human demographics, wealth and education indicators, and governmental interventions.

Given the large heterogeneity between states early on (Fig. 1), we examined correlates of doubling time for only the first seven days after a state reached 25 total cases. We found that population density, flu vaccination rates, and wealth were all correlated with doubling times (Table 1). This suggests that demography might have been more important early in the outbreak, though testing differences between states may also have been important (Fig. S4, Kaashoek & Santillana (2020)).

**Table 1:** Best fitting linear models (according to AIC) and corresponding parameter estimates for the doubling time both early (first 7 days since 25 cases) and for the entire time period.

|  | *Dependent variable:* |  |
| --- | --- | --- |
|  | Early doubling time | Overall doubling time |
| Restrict Restaurants |  | 0.576**<br>(0.119, 1.034) |
| log (Population density) | -0.342***<br>(-0.467, -0.218) | -0.396***<br>(-0.538, -0.253) |
| Vaccination rate | 11.308***<br>(6.914, 15.702) |  |
| GNI per capita | 0.037***<br>(0.021, 0.054) |  |
| Population percent in rural areas |  | 0.016*<br>(0.0003, 0.032) |
| Tightness score |  | -0.024***<br>(-0.040, -0.008) |
| Constant | -12.041***<br>(-16.384, -7.697) | 1.042<br>(-0.545, 2.630) |
| Observations | 50 | 50 |
| R<sup>2</sup> | 0.554 | 0.644 |
| Adjusted R<sup>2</sup> | 0.525 | 0.612 |
| *Note:* |  | *p<0.1; **p<0.05; ***p<0.01 |

We then examined the overall (all days since 25 total cases) doubling times at the state level. Except for population density and percent of population living in rural areas, we found that demography, education, and wealth were poor predictors of the state-level overall doubling times (Fig. 2, Table 1). Therefore, we also examined the correlation between doubling time and state government interventions. We used information collected by Adolph *et al*. (2020) on whether or not a state had implemented a specific action by the first day they had 25 or more cases. We adjusted this number to 150 cases for more severe restrictions like closing all non-essential business or stay at home mandates). We looked at closing of public schools, limiting large gatherings (usually of more than 10 people), restricting business, and stay at home orders (Adolph *et al*. 2020). Of these factors, only restricting businesses, specifically restaurants, was a significant predictor of doubling time (Fig. 3, Table 1). These restrictions were also additive, as states that implemented more actions early had higher doubling times (Fig. 3). The ordering of these restrictions was also fairly consistent between states (Fig. 4). While declaring a state of emergency is an obvious first intervention, closing public schools tend to be implemented at different times across states. More importantly, by the time government restrict businesses and declare a stay-at-home order, all other interventions tend to have already been implemented (Fig. 4, Adolph *et al*. (2020)).

**Figure 2:**
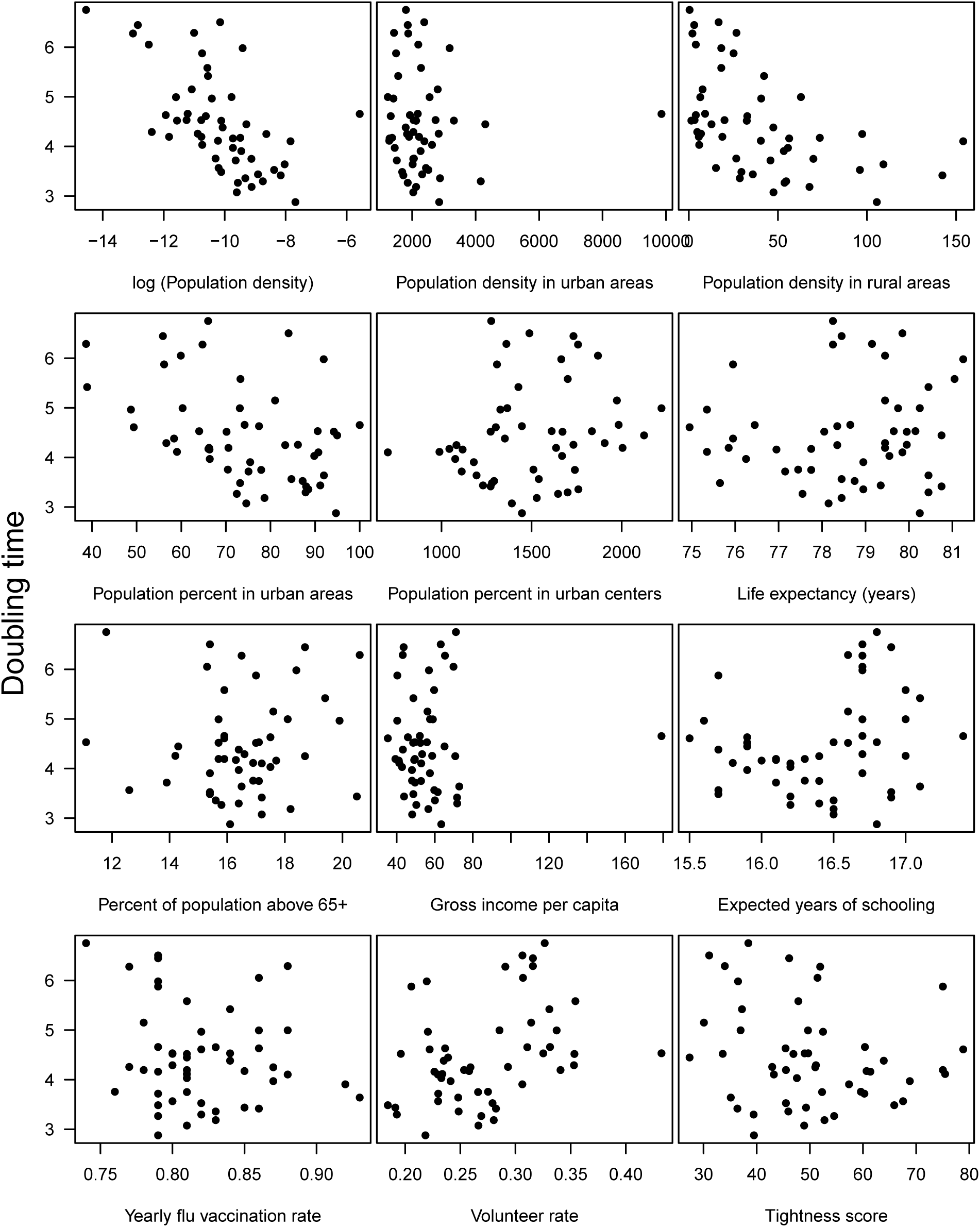
Doubling time (in number of days) versus (a) log (population density), (b) population density in urban areas, (c) population density in rural areas, (d) population percent in urban areas, (e) population percent in urban centers, (f) life expectancy (years), (g) percent of population above age 65, (h) gross income per capita (in 1000s USD), (i) expected years of schooling, (j) yearly flu vaccination rate, (k) volunteer rate, and (l) tightness score.

**Figure 3:**
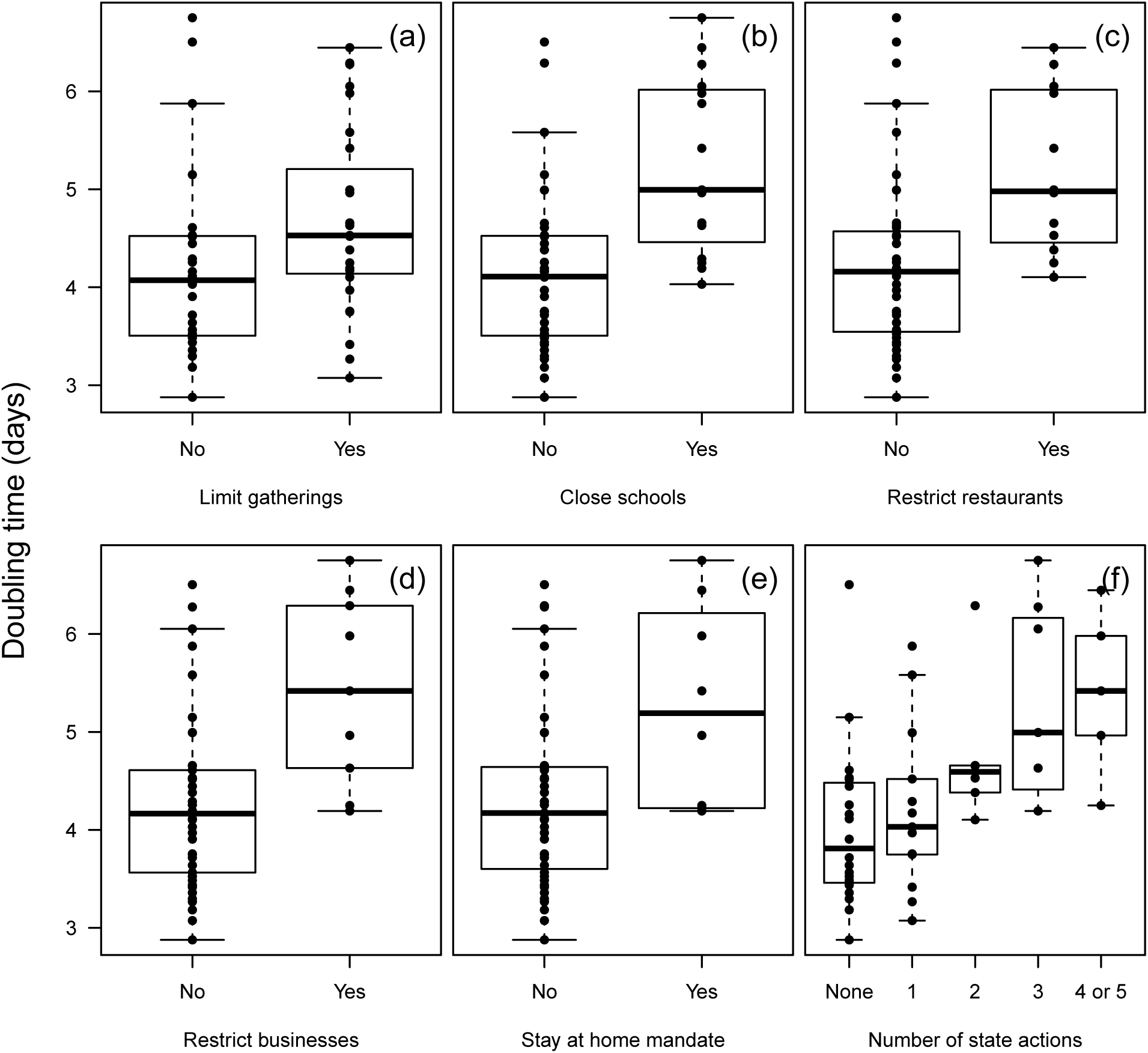
Doubling time (in number of days) across the US states for five different statewide government restrictions: (a) limit gatherings (usually to less than 10 people) by first day of 25 cases, (b) close public schools by first day of 25 cases, (c) restrict restaurants by first day of 25 cases, (d) restrict non-essential businesses by first day of 150 cases, (e) stay at home order by first day of 150 cases, and (f) total number of restrictions before number of cases threshold.

**Figure 4:**
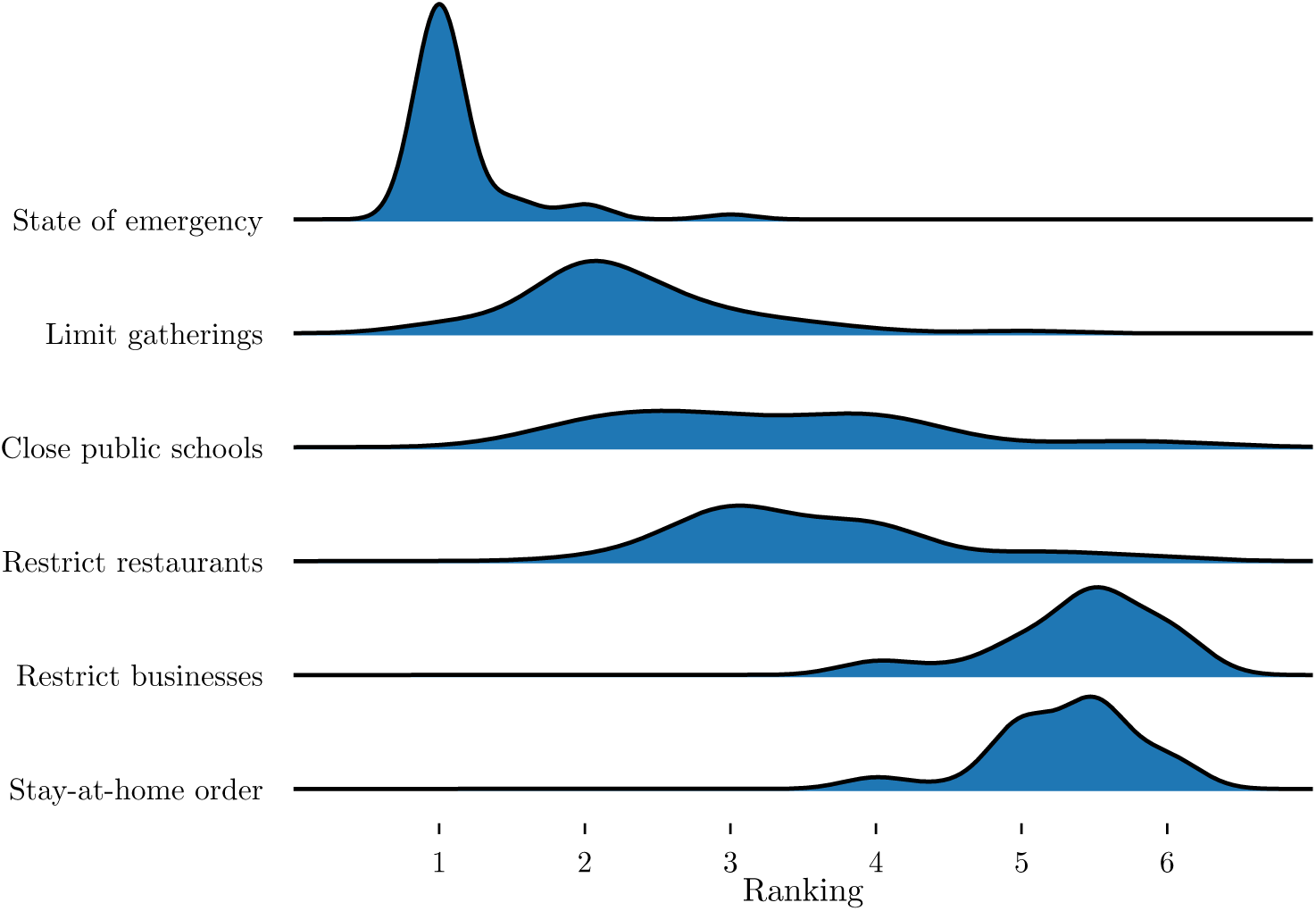
Rank distribution of different interventions. Per state, every intervention is given a rank from 1 to 6 depending on when it was implemented (1 being the first put into place) and ties are given an average rank (e.g. 2.5 for tied 2nd and 3rd rank).

Lastly, after accounting for population density, a state’s “tightness” score was also correlated with doubling time. A state with a high tightness score has “many strongly enforced rules and little tolerance for deviance” (Harrington & Gelfand 2014). We expected that states with highly enforced rules should have higher doubling times compared to “loose” states. Instead, we found that opposite where tight states had low doubling times and faster disease spread. We hypothesis this may be the result of people in tight cultures finding it more difficult to adjust their behavior when new rules are imposed. More work has to be done to understand this relationship.

## Conclusions and Future Work

We found a large degree of heterogeneity in the reported number of COVID-19 cases over time across US states. After state-level government actions were implemented, doubling time was most strongly correlated to restrictions on businesses, in particular restaurants. More detailed work will be needed to understand how these dynamics differ within each state, especially as many government actions started on more local scales.

## Data Availability

All code and corresponding data is freely available at https://github.com/eastonwhite/COVID19_US_States. The original raw data has been compiled by the Johns Hopkins University Center for Systems Science and Engineering at (https://github.com/CSSEGISandData/COVID-19).

## Code availability and acknowledglements

All code and corresponding data is freely available at https://github.com/eastonwhite/COVID19_US_States. The original raw data has been compiled by the Johns Hopkins University Center for Systems Science and Engineering at (https://github.com/CSSEGISandData/COVID-19). L.H.-D. acknowledges support from the National Institutes of Health 1P20 GM125498-01 Centers of Biomedical Research Excellence Award.

### Data sources

- population density and distribution (2010 US Census Bureau https://www.census.gov/programs-surveys/geography/guidance/geo-areas/urban-rural/2010-urban-rural.html)
- percent of population over age 65 (U.S. Census Bureau, Vintage 2018 Population Estimates https://www.prb.org/which-us-states-are-the-oldest/)
- life expectancy, income per capita, and expected years of schooling (Global Data Lab https://globaldatalab.org/shdi/download/2018/indicators/USA/?interpolation=0&extrapolation=0&nearest_real=0&format=csv)
- yearly flu vaccination rate (ChildVaxView CDC https://worldpopulationreview.com/states/vaccination-rates-by-state/)
- volunteer rate (2015 Corporation for National and Community Service data https://www.nationalservice.gov/vcla/state-rankings-volunteer-rate)
- tightness scores Harrington & Gelfand (2014)
- testing rates by state (COVID Tracking Project https://covidtracking.com/)

## Supplemental figures

**Figure S1:**
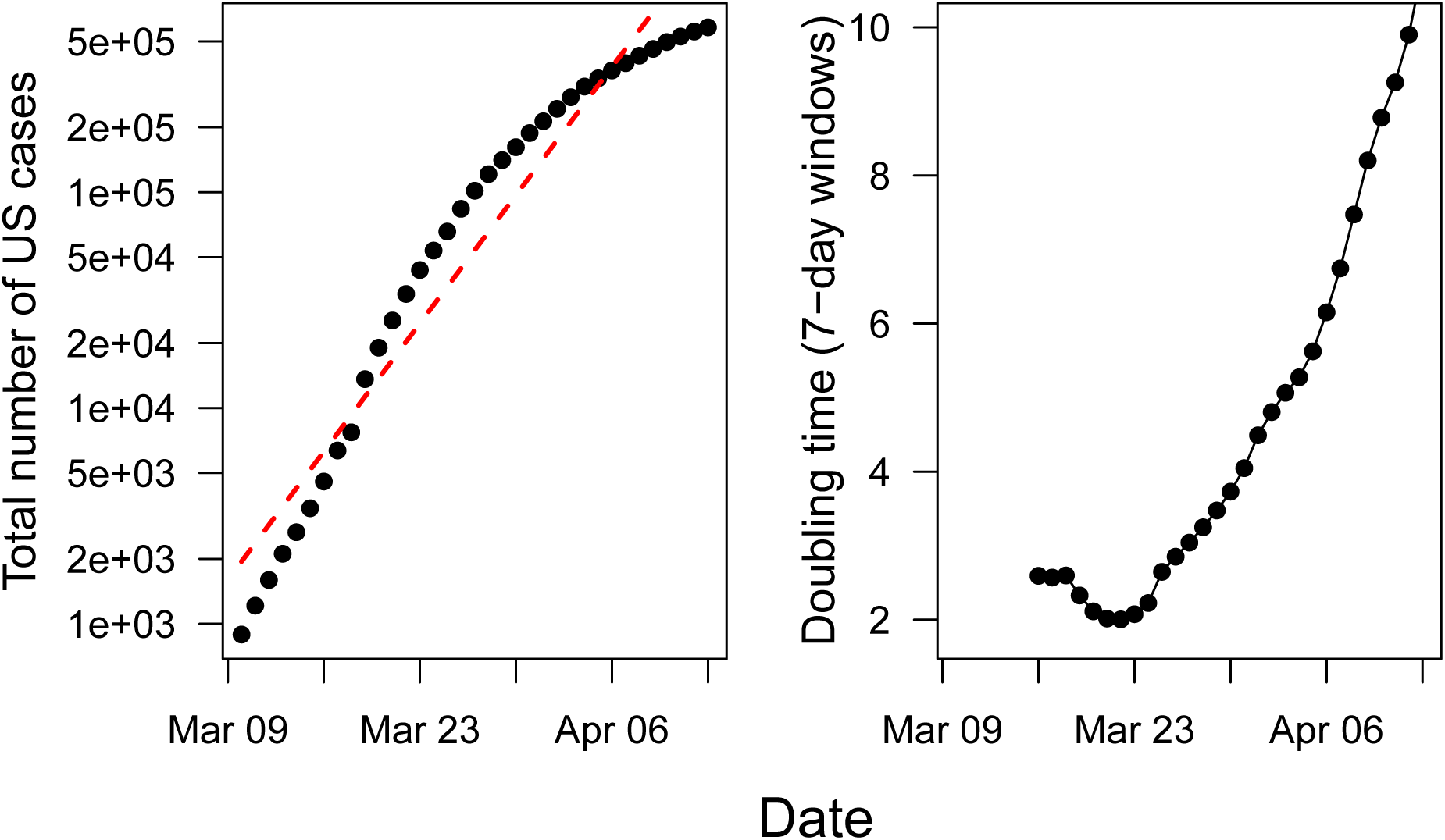
(Left panel) Cases versus time for the whole United States. (Right panel) Log number of cases versus time for the whole United States. The red, dashed line is the line of best fit for all the data and the blue, solid line is the line of best fit since Feburary 29th.

**Figure S2:**
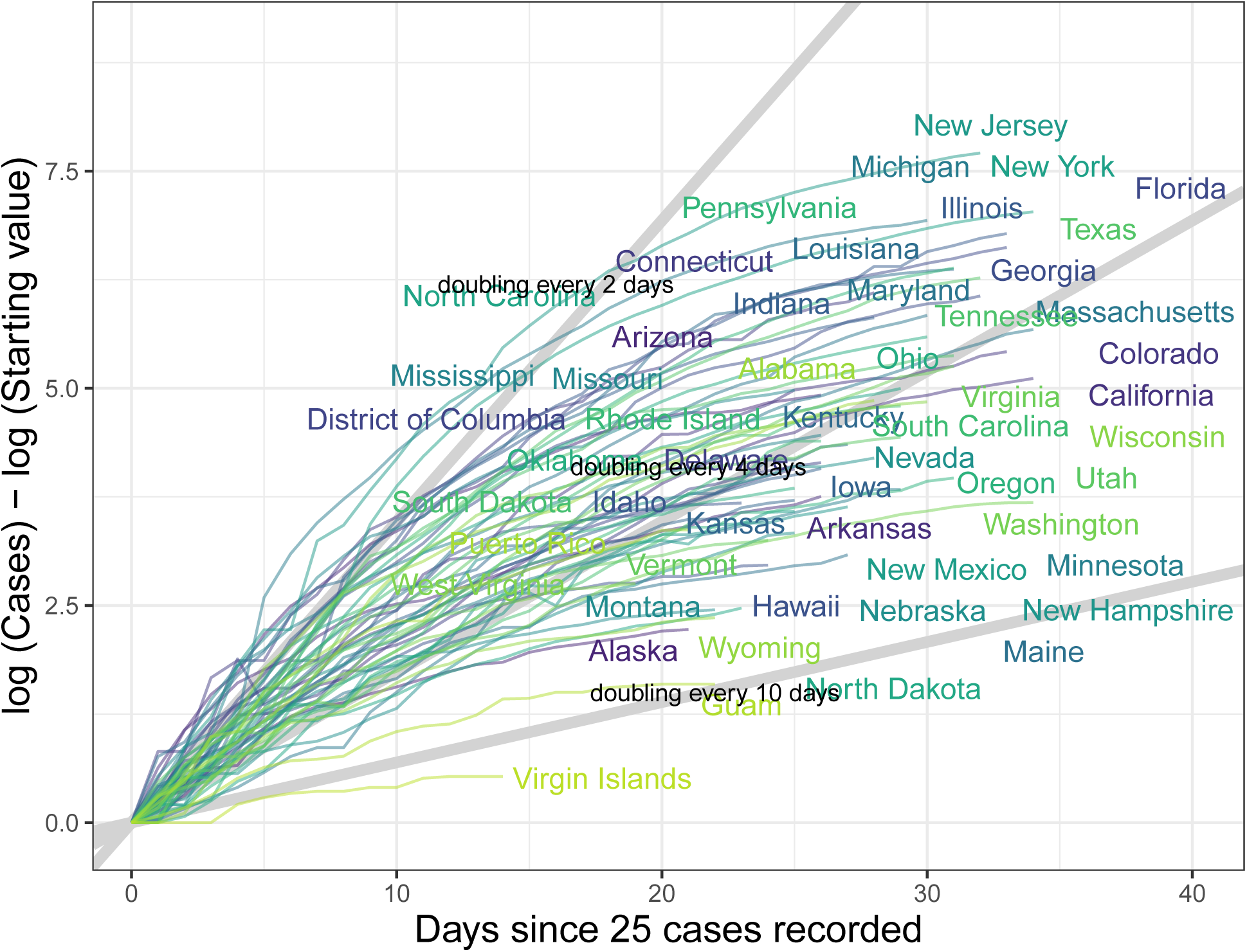
The log number of cases over time for each individual state that recorded more than 25 cases over at least three days. The light grey diagonal lines represent the growth trajectory for doubling times of 2, 4, and 10 days. The log number of the starting value (intial number of cases on first day when at least 25 cases were recorded) had to be subtracted on the y-axis to standarize the graph across states.

**Figure S3:**
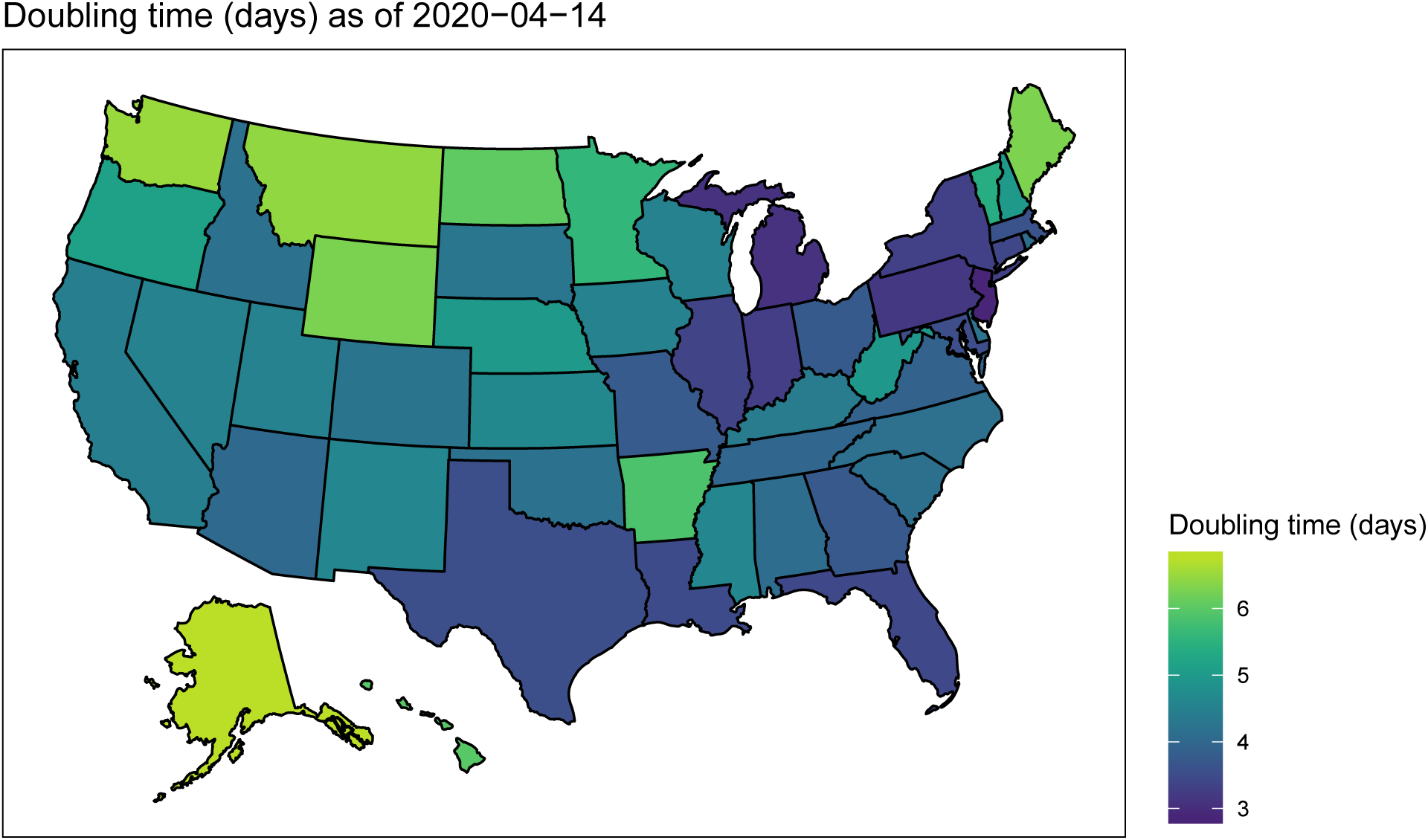
Doubling time (in number of days) across the US states that recorded more than 25 cases over at least three days.

**Figure S4:**
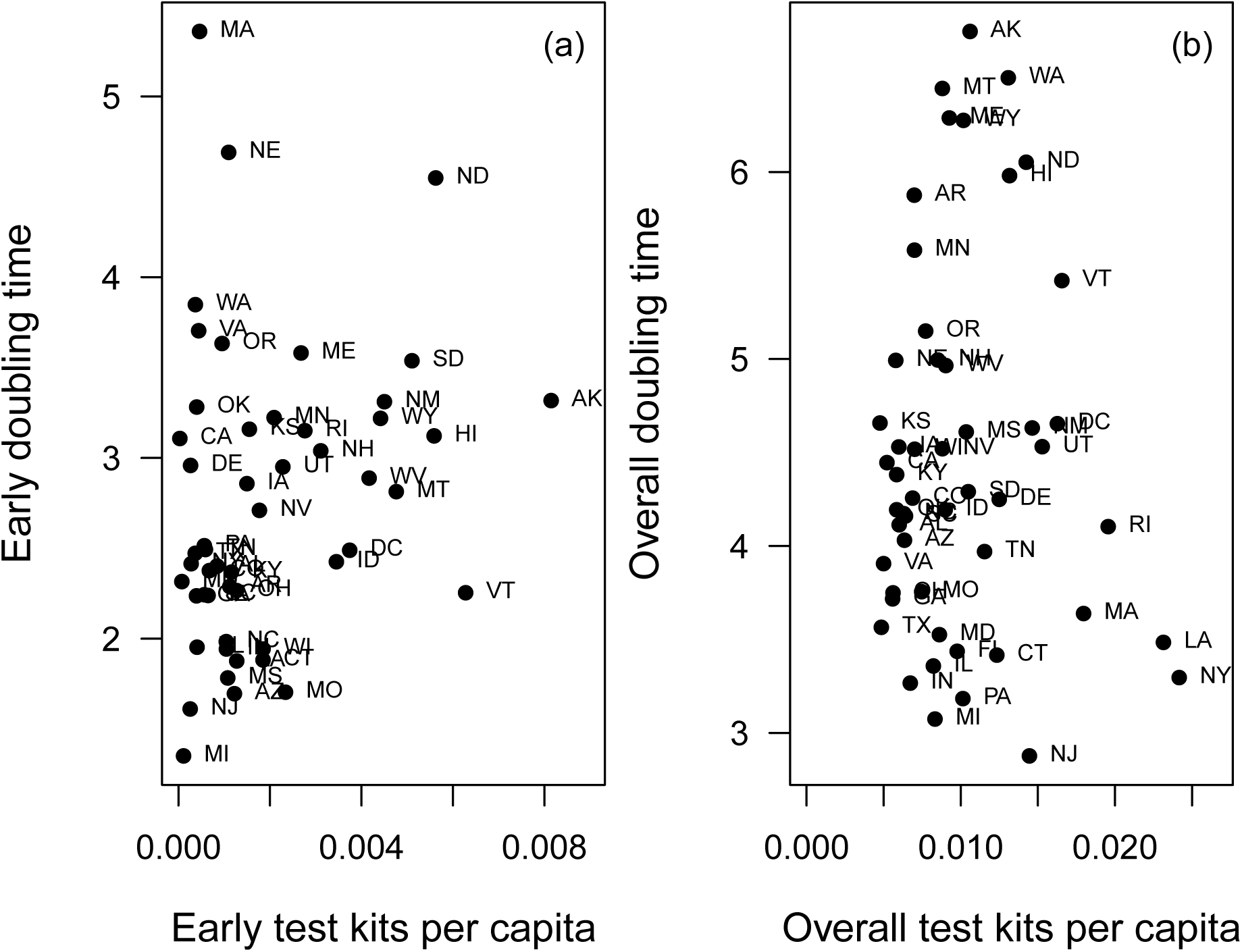
Doubling time (in number of days) for each US state according to their per-capita testing rates (a) early in the outbreak state (within the first week since 25 cases) and (b) for the entire time series.

